# Association Between Coronary Microvascular Parameters and Periprocedural Myocardial Injury in Uncomplicated Elective Percutaneous Coronary Intervention

**DOI:** 10.1101/2025.07.02.25330774

**Authors:** Pedro Jallad, Danilo Ferraz de Oliveira Maksud, Hector M. Garcia-Garcia, Bruno Alves da Mota Rocha, Jean Carlo Mayta Calderon, Roger Renault Godinho, Brunna Pileggi Azevedo Sampaio, Neuza Helena Moreira Lopes, Luis Henrique Wolff Gowdak, Pedro Melo, Antonio Fernando Diniz Freire, Alexandre de Matos Soeiro, Stephanie Itala Rizk, Roberto Kalil Filho, Ludhmila Hajjar, Alexandre Abizaid, Carlos M. Campos

## Abstract

**Background:** Coronary microcirculation is essential for myocardial perfusion and influences clinical outcomes. The angiographic-derived index of microvascular resistance (AMR) is a promising non-invasive tool for assessing microvascular function, but its link to periprocedural myocardial injury (pMI) is unclear.

**Objectives:** To examine coronary flow and microvascular resistance changes during elective PCI and assess the association between post-PCI microvascular dysfunction (measured by AMR) and pMI occurrence.

**Methods:** Patients with stable coronary artery disease (CAD) undergoing elective PCI from June 2021 to December 2023 were included. Coronary physiology was assessed using AMR, quantitative flow ratio (μFR), and coronary flow velocity ratio (CFVR). High-sensitive troponin (hsT) levels were measured post-PCI, with pMI defined by the 4th universal definition.

**Results:** Among 330 patients, pMI occurred in 184 (55.8%). Post-PCI, μFR increased from 0.64 ± 0.21 cm/s to 0.94 ± 0.06 cm/s (p < 0.01), and AMR rose significantly (from 174.92 ± 71.88 to 256.22 ± 55.61 mmHg·s/m, p < 0.01). Microvascular resistance increased in 86.96% of patients. In pMI patients, coronary flow declined (Delta CFVR: - 1.53 ± 5.38 vs. 0.26 ± 4.95, p = 0.03) and AMR was significantly higher (265.4 ± 56.4 mmHg*s/m vs 244.7 ± 52.4 mmHg*s/m, p < 0.01). Microvascular dysfunction was more common in pMI patients (56.5% vs. 39.7%, p < 0.01), especially those with reduced flow velocity (50.0% vs. 19.1%, p < 0.01).

**Conclusion:** This study shows that increased microvascular resistance after PCI— indicated by elevated AMR and reduced RVR—is associated with a higher risk of pMI, while baseline values are not. Despite successful epicardial revascularization, these indices may help guide PCI and evaluate outcomes.

## 1 Introduction

Myocardial injury in the context of coronary disease is primarily characterized by an elevation of troponin above the 99th percentile upper reference limit, often occurring after percutaneous coronary intervention (PCI) or acute coronary syndromes. Its incidence varies but is commonly observed following PCI, with studies reporting rates ranging from 5% to 30%, depending on the definition used and patient population (1). The mechanisms involved includes microvascular dysfunction, distal embolization, side branch occlusion, and ischemia-reperfusion injury (2). Additional contributing factors include endothelial dysfunction, coronary plaque disruption, and procedural complications leading to transient myocardial ischemia (3). While myocardial injury does not always progress to infarction, its presence is associated with worse cardiovascular outcomes. Identifying the mechanisms underlying microvascular impairment following PCI is therefore critical to improving patient prognosis (4).

In recent years, novel physiological indices derived from coronary angiography, including Murray law-based quantitative flow ratio (μFR), coronary flow velocity ratio (CFVR), and angiographic-derived microcirculatory resistance (AMR), have emerged as valuable tools for assessing coronary physiology (5–7). These indices provide insights into both epicardial and microvascular function without requiring invasive pressure-wire measurements. Studies suggest that post-PCI microvascular dysfunction, as indicated by abnormal values in these indices, may be associated with an increased risk of pMI, highlighting the need for further investigation. (8–9) The present study aims to investigate angio-derived coronary flow and microvascular resistance during elective PCI, and to explore the relationship between post-PCI microvascular dysfunction and the occurrence of periprocedural myocardial injury (pMI).

## 2 Methods Study design

The study population was obtained through retrospective analysis of electronic medical records of the Heart Institute of the School of Medicine, University of São Paulo (InCor/FMUSP) between June 2021 and December 2023. Consecutive patients over 18 years old who underwent uncomplicated successful elective PCI for stable coronary artery disease (CAD) were eligible for inclusion. PCI success was defined as residual stenosis <30% associated with Thrombolysis in Myocardial Infarction (TIMI) III coronary flow, without side branch occlusion (10).

Exclusion criteria were based on conditions that could affect the analysis of μFR or alter troponin levels: ostial lesions, surgical graft lesions, bifurcation requiring a two-stent technique, chronic total occlusion, occurrence of slow flow/no-reflow during the procedure, coronary perforation, coronary dissection, use of a left ventricular assist device during PCI, and poor image quality. Baseline characteristics and laboratory measurements were collected in a dedicated study database (REDCap – Research Eletronic Data Capture). The Medical Ethics Committee approved this retrospective evaluation and waived the need for written informed consent (CAAE number 01798912.1.0000.0181).

### Angiographic Analysis – μFR, CFVR and AMR

The angiographic analysis was performed using μFR software (AngioPlus Core, version V3, Shanghai Pulse Medical Technology Inc., Shanghai, China). Analysis was made by an experienced, certified analyst in an independent academic laboratory, blinded to procedural invasive data or clinical outcomes. The analyst was blinded to the results of laboratory tests. The detailed methodology for calculating μFR (11) consists of selecting the optimal angiographic view with minimal vessel overlap, the lumen contour of the investigated coronary artery is automatically delineated, while the contrast flow velocity is derived from the centerline length of the vessel divided by contrast filling time, and then converted into hyperemic flow velocity (12).

Subsequently, a frame with good contrast fill and full exposure of the lumen contour is selected as the analysis frame, and the boundaries of the vessel lumen and main side branches are automatically delineated. The reference diameter of the vessel is then reconstructed, considering the degradation phenomenon at bifurcations based on Murray’s fractal bifurcation law (13,14). Finally, modeling of hyperemic flow velocity based on the contrast flow velocity and calculation of pressure drop based on fluid dynamics Equation (15), assuming a blood density of 1,060 kg/m³ and viscosity of 0.0035 kg/(m.s). Specifically, the pressure loss caused by the frictional loss along the lesion entrance and stenotic segment as well the inertial loss stemming from the sudden expansion of the flow as it emerged from the stenosis were calculated, based on the stenosis geometry and hyperemic flow rate. In the end, μFR is available in both, main and side branch, without the need for aortic pressure **(Figure 1).**

**Figure 1.**
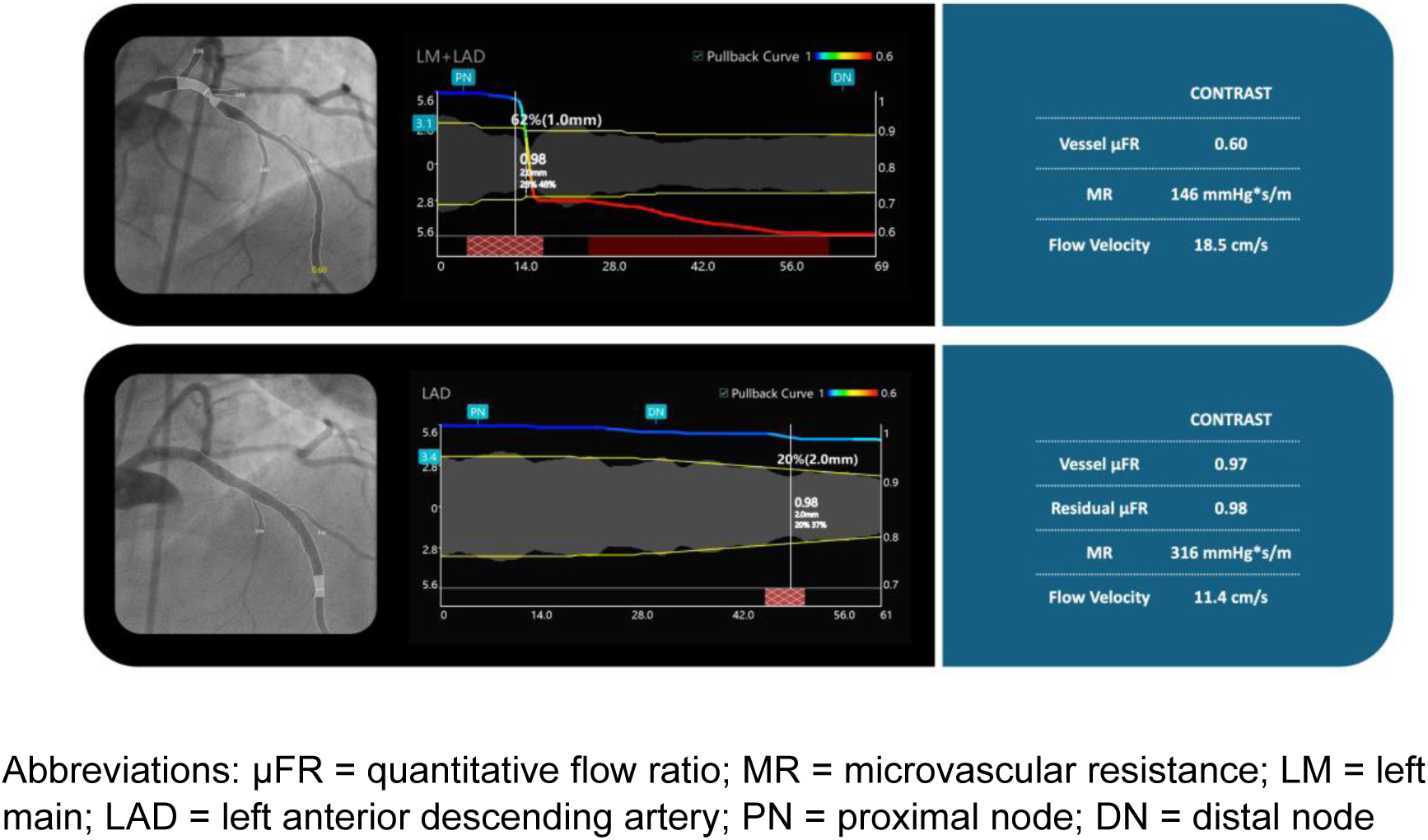
Quantitative Coronary Physiology Assessment in the Left Main and Left anterior descending artery segments. Panels show coronary angiography images, physiological pullback curves, and corresponding quantitative metrics from contrast-based microvascular resistance (MR) assessment in the left anterior descending artery (LAD) before (top) and after (bottom) percutaneous coronary intervention (PCI). Top panel (Pre-PCI): Significant epicardial stenosis is observed with a vessel μFR of 0.60. The microvascular resistance is 146 mmHg*s/m with a Flow Velocity of 18.5 cm/s. Bottom panel (Post-PCI): Following stent implantation, vessel μFR improves to 0.97 and residual μFR to 0.98, confirming effective epicardial revascularization. However, MR increases to 316 mmHg·s/m and Flow Velocity decreases to 11.4 cm/s, suggesting elevated microvascular resistance post-PCI despite anatomical and functional epicardial success.

The AMR is an index used to estimate the resistance to blood flow in the microcirculation. Through the angiogram, μFR software estimates both the CFVR (unit: cm/s) and pressure gradients across different sections of the arteries. Once the flow and pressure data are computed, the machine calculates the AMR (unit: mmHg*s/m), which is defined as: ΔP/CFVR, being ΔP the pressure gradient across the microcirculation **(Figure 1).**

### Clinical Outcomes Measures

At least three hsT measurements every 8 hours after PCI were collected. If a diagnosis of pMI occurred, the patient had subsequent collections until a descending curve is observed. Periprocedural injury was defined as the stablished 4^th^ universal definition, with a threshold for pMI of >5x the hsT reference value. AMR > 250 mmHg*s/ was defined as coronary microvascular disfunction (CMD) (16). A reduced velocity ratio (RVR) was defined as delta CFVR (difference between post and pre PCI) ≤-2.6 cm/s, obtained from Youden Index of delta CFVR’s ROC curve to predict pMI.

## Data Analysis

Continuous variables were expressed as means and standard deviations or medians and interquartile ranges (IQRs), according to the sample distribution pattern. We used the Shapiro–Wilk test to assess distribution. Comparisons were conducted using Kruskal–Wallis for non-parametric one-way anova test or Mann–Whitney for non-parametric t test. Categorical variables were expressed as percentages and compared using the χ 2 test. The programs used for the statistical analysis were GraphPad Prism Statistical Software version 10.2.2 and RStudio 2022.12.0 for Windows, version 22.0.

## 3 Results

### Study population baseline characteristics

From June 2021 until December 2023, 422 patients underwent elective successful PCI for stable CAD at Heart Institute of University of São Paulo (HC-FMUSP). **Figure 2** illustrates the flowchart of inclusion and exclusion of patients.

**Figure 2.**
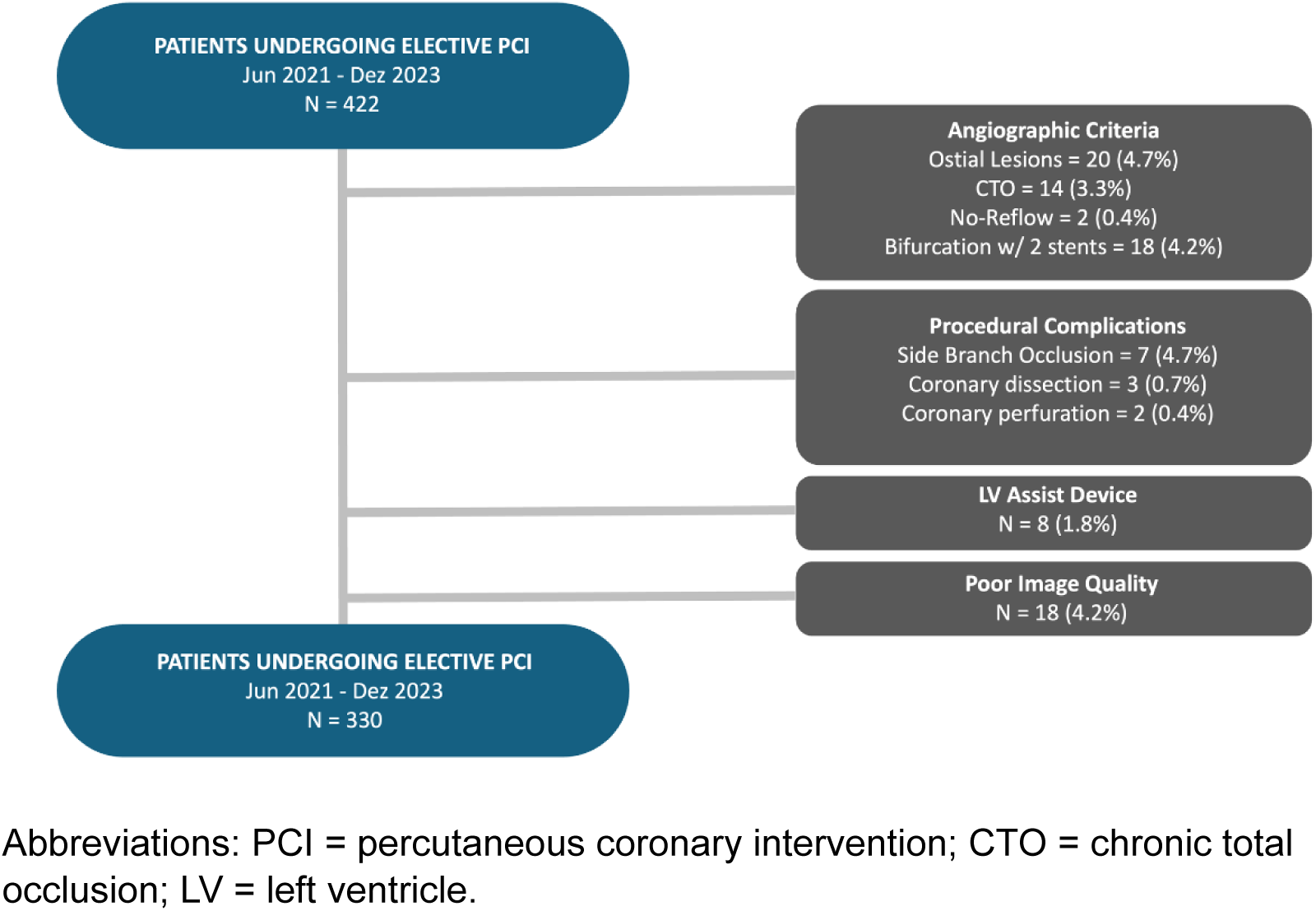
Study population and exclusion criteria for included patients. Flowchart illustrating the selection of patients undergoing elective percutaneous coronary intervention (PCI) between June 2021 and December 2023. From an initial cohort of 422 patients, 92 were excluded based on predefined criteria: angiographic criteria (ostial lesions, chronic total occlusion (CTO), no-reflow phenomenon, or bifurcation treated with two stents), procedural complications (side branch occlusion, coronary dissection, or perforation), use of left ventricular assist devices, or poor image quality. The final study population comprised 330 patients.

Baseline clinical characteristics are shown in **Table 1**. Overall, the median age was 65 years (Interquartile Range [IQR], 58-73 years), male sex 179 patients (59.6%), diabetes mellitus was present in 52.4% and the mean left ventricular ejection fraction (LVEF) was 55.5% ± 10.1.

**Table 1.** Baseline Clinical Characteristics.

|  | Overall<br>N=330 | No PMI<br>N=146 | PMI<br>N=184 | P |
| --- | --- | --- | --- | --- |
| Age, Years (25th-75th percentil) | 65.0 (58.1-73.2) | 64.0 (61.2-69.3) | 65.0 (60.0-72.2) | 0.64 |
| Male sex, n (%) | 179.0 (54.2) | 82.0 (56.1) | 97.0 (52.7) | 0.81 |
| Hypertension, n (%) | 287.0 (86.9) | 135.0 (92.4) | 152.0 (82.6) | 0.74 |
| Dyslipidemia, n (%) | 241.0 (73.1) | 112.0 (76.7) | 129.0 (70.1) | 0.67 |
| DM, n (%) | 173.0 (52.4) | 81.0 (55.4) | 92.0 (50.0) | 0.68 |
| Obesity, n (%) | 102.0 (30.9) | 46.0 (31.5) | 56.0 (30.4) | 0.75 |
| Prior MI, n (%) | 129.0 (39.0) | 49.0 (33.5) | 70.0 (38.0) | 0.45 |
| Angina CCS Class III/IV, n (%) | 239.0 (72.4) | 107 (73.3) | 132 (71.7) | 0.38 |
| COPD, n (%) | 65.0 (19.7) | 30 (20.5) | 35 (19.0) | 0.61 |
| History of câncer (%) | 22 (6.6) | 8 (5.4) | 14 (7.6) | 0.08 |
| PAD (%) | 38 (11.5) | 12 (8.2) | 26 (14.1) | 0.04 |
| Prior coronary angioplasty, n (%) | 98.0 (29.7) | 42.0 (28.7) | 56.0 (30.4) | 0.88 |
| LVEF (%) ± SD | 55.5 ± 10.1 | 56.2 ± 10.0 | 54.7 ± 10.8 | 0.92 |
| CrCl (ml/min) ± SD | 64.6 ± 18.8 | 65.6 ± 18.5 | 65.6 ± 16.7 | 0.79 |
| Troponin/URL (25th-75th) | 6.1 (1.9 – 20.9) | 1.7 (0.8 – 3.0) | 18.1 (8.2 – 37.9) | <0.01 |

Abbreviations: PMI = periprocedural myocardial injury; DM = diabetes mellitus; MI = myocardial infarction; CCS = Canadian Cardiovascular Society; COPD = chronic obstructive pulmonary disease; PAD = peripheral artery disease; LVEF = left ventricular ejection fraction; CrCl = creatinine clearance; URL = upper reference limit; SD = standard deviation.

The mean peak of hsT/URL post PCI was 6.14 (IQR 1.9 – 20.9) and pMI occurred in 184 patients (55.8%). Most of baseline characteristics did not differ between patients with or without pMI, except for peripheral artery disease with a significantly higher prevalence in patients with pMI (14.1% vs. 8.2%, P = 0.04) **(Table 1)**.

### Baseline angiography and procedural characteristics

The angiographic and procedural characteristics of the study population were comparable across the subgroups without pMI (n=146) and with pMI (n=184). There were no statistically significant differences in the distribution of target vessels, stent-related parameters, and maximum stenosis diameter **(Table 2).**

**Table 2.** Angiographic and Procedural Characteristics.

|  | Baseline (330) | No pMI (146) | pMI (184) | P |
| --- | --- | --- | --- | --- |
| Target vessel |  |  |  |  |
| Left Main PCI, n (%) | 20.0 (6.1) | 9.0 (6.1) | 11.0 (5.9) | 0.46 |
| LAD, n (%) | 117.0 (35.4) | 52.0 (35.6) | 65.0 (35.3) | 0.29 |
| Circumflex, n (%) | 81.0 (24.5) | 38.0 (26.0) | 43.0 (23.3) | 0.37 |
| Right coronary PCI, n (%) | 112.0 (33.9) | 47.0 (32.2) | 65.0 (35.3) | 0.55 |
| Mean stent diameter (mm) ± SD | 3.1 ± 0.4 | 3.0 ± 0.4 | 3.2 ± 0.3 | 0.48 |
| Minimum Lumen Diameter (mm) ± SD | 1.1 ± 0.3 | 1.1 ± 0.1 | 1.1 ± 0.4 | 0.19 |
| Maximum Stenosis Diameter (%) ± SD | 78.1 ± 12.2 | 76.2 ± 12.0 | 77.7 ± 12.6 | 0.63 |
| Proximal diameter (mm) ± SD | 2.7 ± 0.6 | 2.7 ± 0.4 | 2.6 ± 0.5 | 0.58 |
| Distal diameter (mm) ± SD | 2.3 ± 0.4 | 2.40 ± 0.4 | 2.3 ± 0.4 | 0.49 |
| Pre dilatation balloon diameter (mm) ± SD | 3.6 ± 0.1 | 3.5 ± 0.3 | 3.6 ± 0.2 | 0.81 |
| Post dilatation balloon diameter (mm) ± SD | 3.3 ± 0.4 | 2.3 ± 0.3 | 2.4 ± 0.2 | 0.74 |
| Radial Access, n (%) | 267.0 (80.9) | 117.0 (80.1) | 150.0 (81.5) | 0.43 |
| Femoral Access, n (%) | 63.0 (19.1) | 29.0 (19.9) | 34.0 (18.5) | 0.41 |

Abbreviations: left anterior descending (LAD); PCI = percutaneous coronary intervention; LAD = left anterior descending artery; SD = standard deviation; pMI = periprocedural myocardial injury.

### Coronary Physiology in overall population

Figure 3 shows the overall coronary physiology pre and post-PCI. Pre-PCI μFR was 0.64 ± 0.21 cm/s, while post-PCI μFR increased to 0.94 ± 0.06 cm/s (P < 0.01).

**Figure 3.**
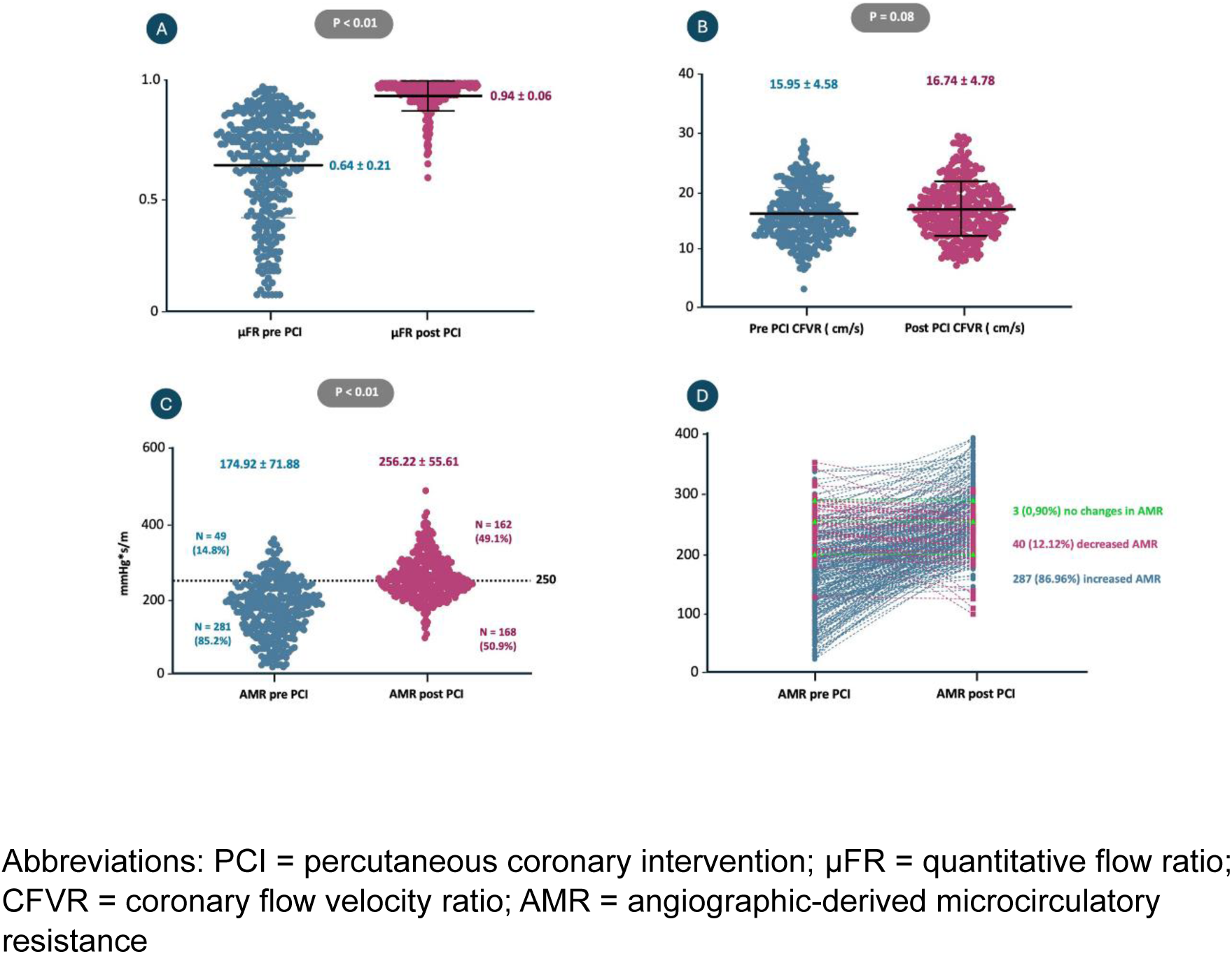
Changes in coronary physiology before and after elective PCI. (A) Vessel-level contrast-based μFR significantly increased after PCI (from 0.64 ± 0.21 to 0.94 ± 0.06, *P*<0.01), indicating successful epicardial revascularization. (B) Coronary flow velocity ratio (CFVR) showed a non-significant increase post-PCI (from 15.95 ± 4.58 to 16.74 ± 4.78 cm/s, *P* = 0.08). (C) angiographic-derived microcirculatory resistance (AMR) increased significantly after PCI (from 174.92 ± 71.88 to 256.22 ± 55.61 mmHg*s/m, *P*<0.01). Based on a post-PCI AMR threshold of 250 mmHg*s/m, 49.1% of patients showed elevated AMR. (D) Individual patient trajectories of AMR before and after PCI demonstrate that 86.96% (n = 287) had increased AMR post-procedure, while only 12.12% (n = 40) had decreased AMR, and 0.90% (n = 3) showed no change.

After stent implantation it was observed a significant rise in microvascular resistance (AMR pre-PCI:174.92 ± 71.88 mmHg*s/m and post-PCI: 256.22 ± 55.61 mmHg*s/m; P < 0.01) (Figure 3C). In total, 86.96% (n = 287) of patients experienced an increase in AMR following the procedure, while 12.12% (n = 40) exhibited a decrease, and 0.90% (n = 3) maintained stable AMR values (Figure 3D). In relation to CFVR, there was no substantial difference on pre and post PCI values (P = 0.08) (Figure 3B).

### Periprocedural MI and Coronary Physiologic Indexes

There were significant differences between patients experiencing pMI, particularly in coronary flow and microvascular function, as shown in **Table 3** and Figure 4. Patients presenting with pMI exhibited a decline in coronary flow (Delta CFVR:-1.53 ± 5.38), while the non-pMI group demonstrated an increase in coronary flow (Delta CFVR: 0.26 ± 4.95; *P* = 0.03). Importantly, visual assessment confirmed that all patients included in this analysis achieved a final TIMI flow grade III.

**Figure 4.**
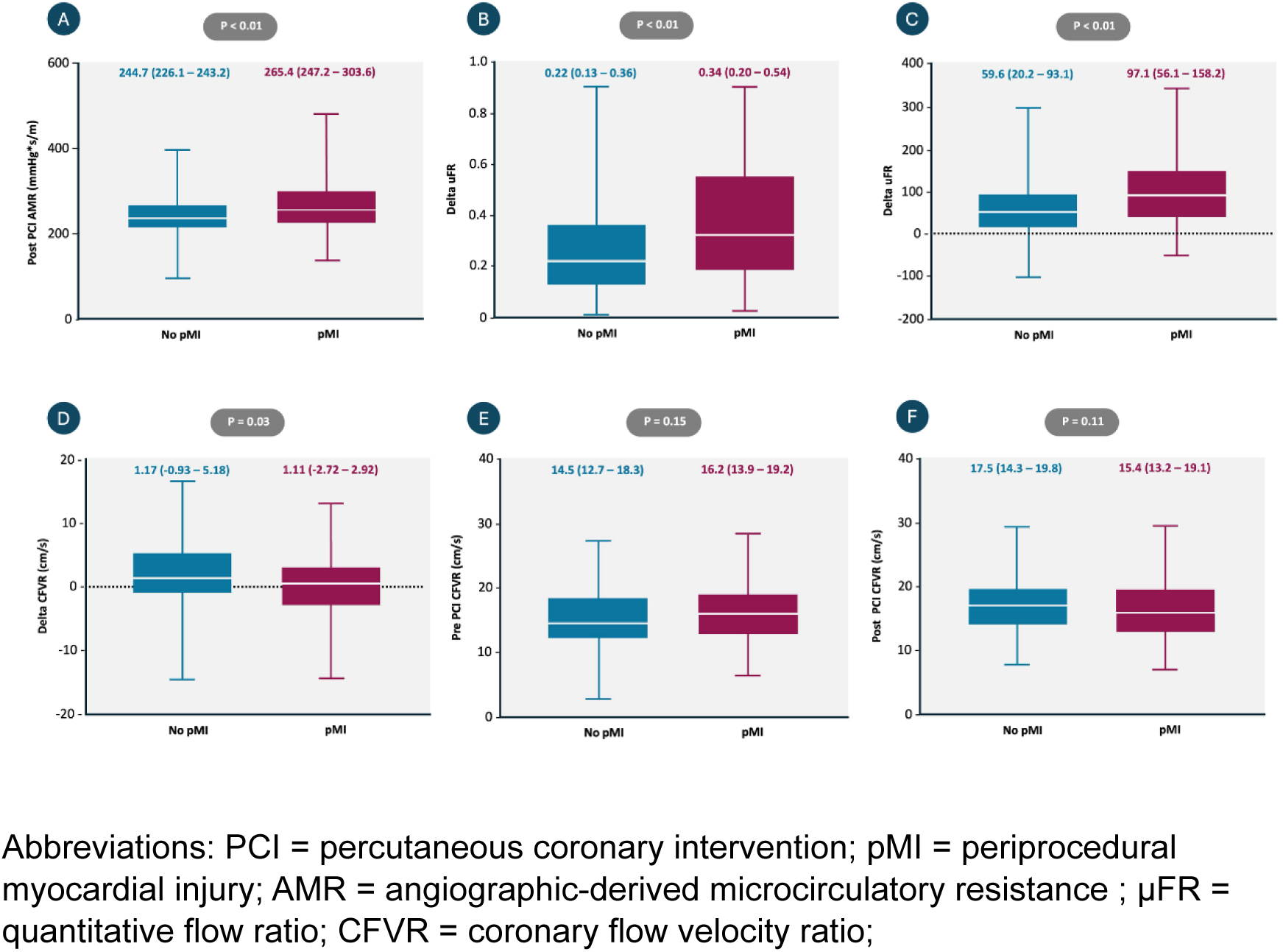
Comparison of coronary physiological parameters between patients with and without periprocedural myocardial injury (pMI). (A) Post-PCI angiographic microvascular resistance (AMR) was significantly higher in patients with pMI compared to those without (265.4 vs. 244.7 mmHg*s/m; P<0.01). (B) Delta μFR (post-PCI minus pre-PCI) was significantly greater in the pMI group (0.34 vs. 0.22; P<0.01). (C) Delta AMR (post-PCI minus pre-PCI) was also significantly greater in patients with pMI (97.1 vs. 59.6 mmHg*s/m; P<0.01). (D) Delta CFVR showed a significant decrease in the pMI group compared to no pMI (1.11 vs. 1.17; P = 0.03). (E) Pre-PCI CFVR did not differ significantly between groups (16.2 vs. 14.5 cm/s; P = 0.15). (F) Post-PCI CFVR also showed no significant difference (15.4 vs. 17.5 cm/s; P = 0.11). Data are presented as median [interquartile range].

**Table 3.**
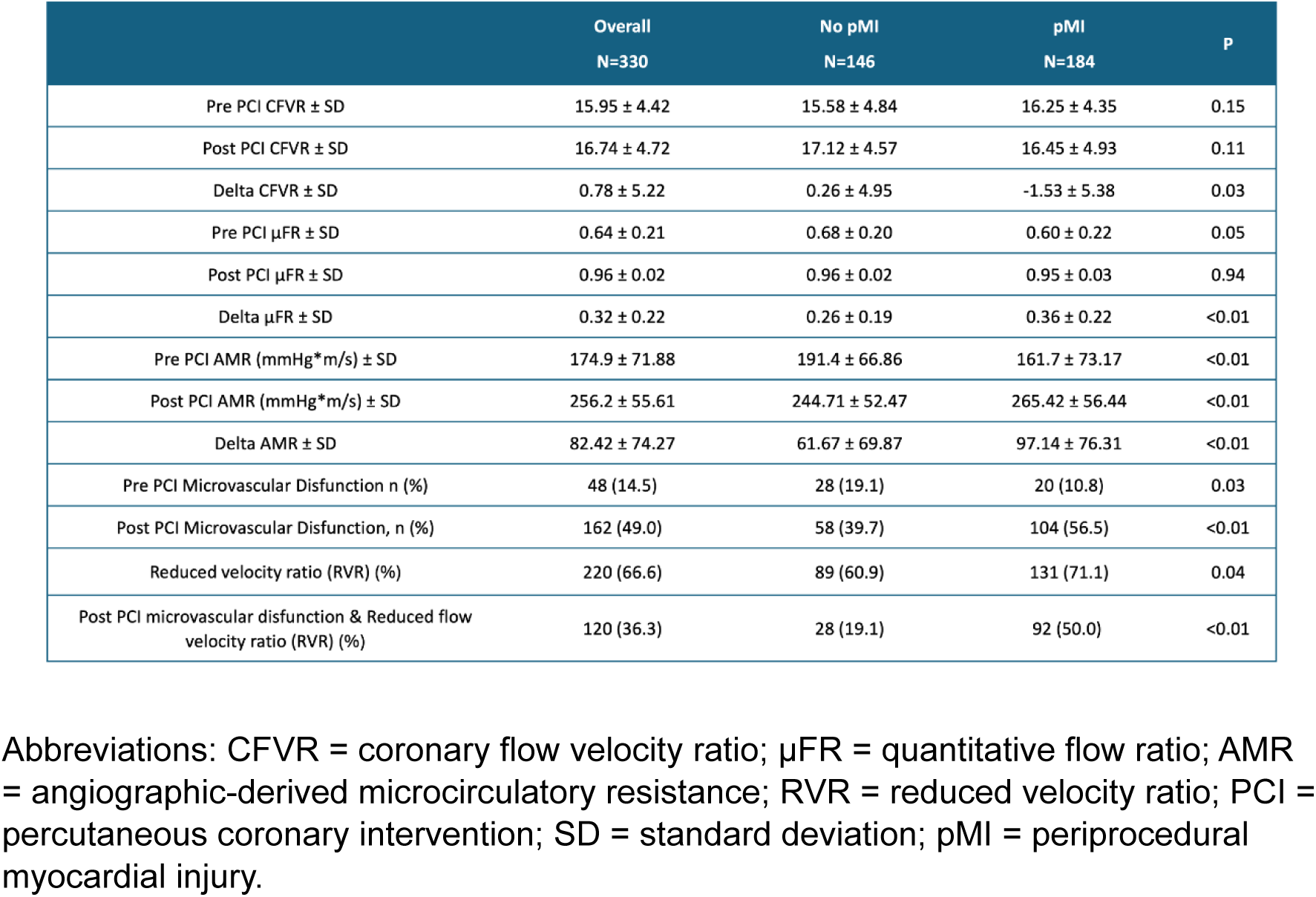
Angiographic-Derived Physiologic Indexes.

|  | Overall<br>N=330 | No pMI<br>N=146 | pMI<br>N=184 | P |
| --- | --- | --- | --- | --- |
| Pre PCI CFVR ± SD | 15.95 ± 4.42 | 15.58 ± 4.84 | 16.25 ± 4.35 | 0.15 |
| Post PCI CFVR ± SD | 16.74 ± 4.72 | 17.12 ± 4.57 | 16.45 ± 4.93 | 0.11 |
| Delta CFVR ± SD | 0.78 ± 5.22 | 0.26 ± 4.95 | -1.53 ± 5.38 | 0.03 |
| Pre PCI μFR ± SD | 0.64 ± 0.21 | 0.68 ± 0.20 | 0.60 ± 0.22 | 0.05 |
| Post PCI μFR ± SD | 0.96 ± 0.02 | 0.96 ± 0.02 | 0.95 ± 0.03 | 0.94 |
| Delta μFR ± SD | 0.32 ± 0.22 | 0.26 ± 0.19 | 0.36 ± 0.22 | <0.01 |
| Pre PCI AMR (mmHg*m/s) ± SD | 174.9 ± 71.88 | 191.4 ± 66.86 | 161.7 ± 73.17 | <0.01 |
| Post PCI AMR (mmHg*m/s) ± SD | 256.2 ± 55.61 | 244.71 ± 52.47 | 265.42 ± 56.44 | <0.01 |
| Delta AMR ± SD | 82.42 ± 74.27 | 61.67 ± 69.87 | 97.14 ± 76.31 | <0.01 |
| Pre PCI Microvascular Dysfunction n (%) | 48 (14.5) | 28 (19.1) | 20 (10.8) | 0.03 |
| Post PCI Microvascular Dysfunction, n (%) | 162 (49.0) | 58 (39.7) | 104 (56.5) | <0.01 |
| Reduced velocity ratio (RVR) (%) | 220 (66.6) | 89 (60.9) | 131 (71.1) | 0.04 |
| Post PCI microvascular dysfunction & Reduced flow velocity ratio (RVR) (%) | 120 (36.3) | 28 (19.1) | 92 (50.0) | <0.01 |

Abbreviations: CFVR = coronary flow velocity ratio; μFR = quantitative flow ratio; AMR = angiographic-derived microcirculatory resistance; RVR = reduced velocity ratio; PCI = percutaneous coronary intervention; SD = standard deviation; pMI = periprocedural myocardial injury.

As shown in **Table 3**, at baseline, patients without pMI had nearly twice the degree of microvascular dysfunction (19.1% vs. 10.8%; P=0.03). However, a marked increase in post-PCI AMR was observed in the pMI group (*P* < 0.01 for post-PCI AMR and Delta AMR). Consequently, post-PCI microvascular dysfunction became more prevalent in the pMI group compared to the non-pMI group (56.5% vs. 39.7%; *P* < 0.01).

Additionally, patients with pMI more frequently demonstrated a combination of RVR and microvascular disfunction (50.0% vs. 19.1%, P < 0.01), as show in Figure 5.

**Figure 5.**
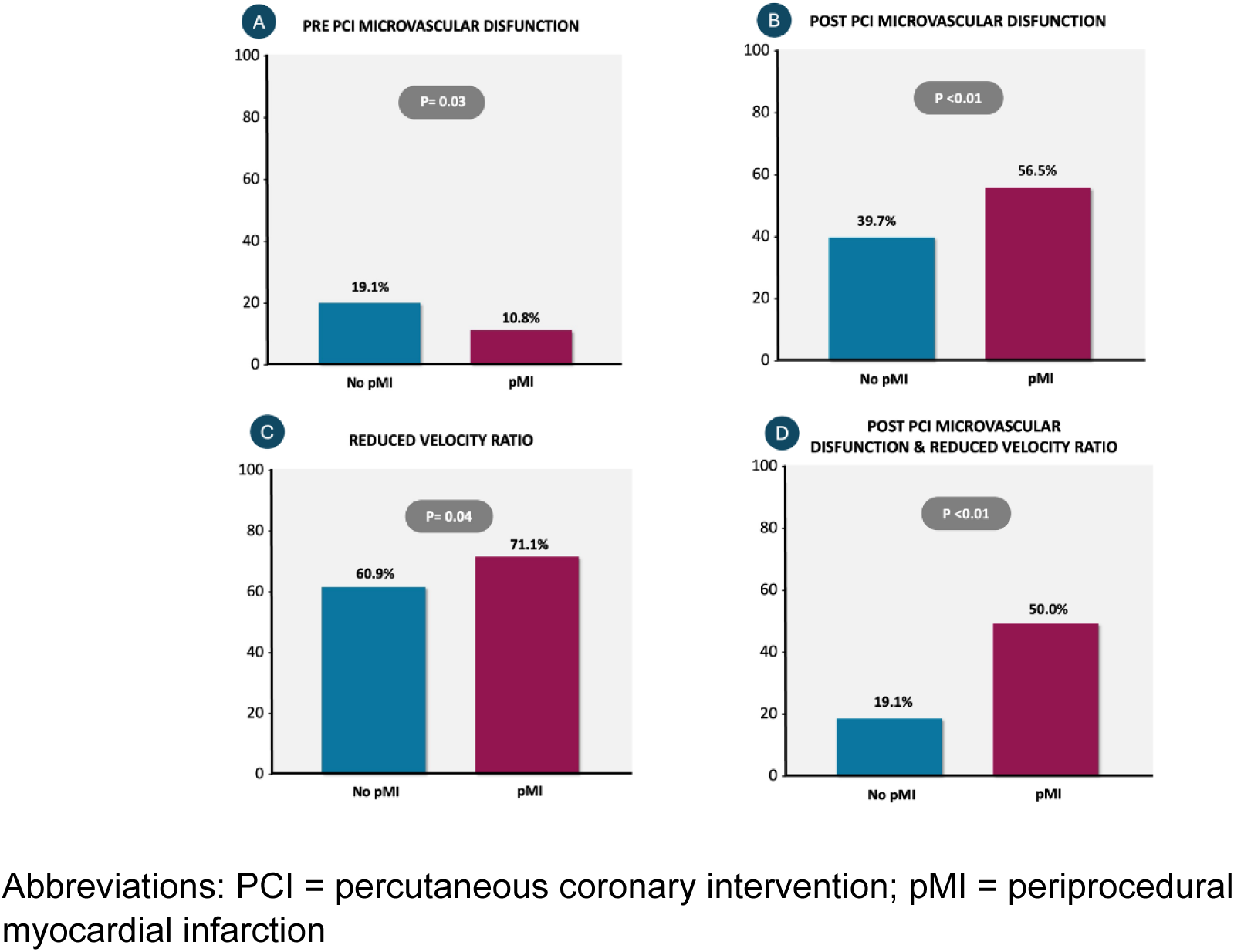
Association between microvascular dysfunction, reduced velocity ratio, and the occurrence of periprocedural myocardial injury (pMI). (A) Pre-PCI microvascular dysfunction (defined by AMR >250 mmHg*s/m) was more frequent in patients without pMI compared to those with pMI (19.1% vs. 10.8%; *P* = 0.03). (B) In contrast, post-PCI microvascular dysfunction occurred significantly more often in patients with pMI (56.5%) than those without (39.7%; *P*<0.01). (C) A reduced velocity ratio (RVR) was more prevalent in the pMI group (71.1% vs. 60.9%; *P* = 0.04). (D) The combination of post-PCI microvascular dysfunction and reduced velocity ratio was significantly more common among patients with pMI (50.0%) than those without (19.1%; *P*<0.01), suggesting a strong association with adverse outcomes.

## 4 Discussion

The main findings of the present study can be summarized as follows: (1) after an uncomplicated, successful elective percutaneous coronary intervention, coronary flow velocity accelerates, and microvascular resistance increases; (2) microvascular dysfunction is a frequent finding, with an incidence of 49%; (3) the occurrence of periprocedural myocardial injury (pMI) is often associated with coronary flow deceleration and elevated microvascular resistance.

Our analysis of coronary physiology before and after PCI demonstrates a significant improvement in coronary flow following successful revascularization. Pre-PCI, μFR was 0.64 ± 0.21 cm/s, which markedly improved post-PCI to 0.94 ± 0.06 cm/s, indicating a substantial reduction in epicardial stenosis. Subsequently, we observed an overall increase in coronary flow velocity and absolute AMR.

Invasive methods for evaluating epicardial and CMD have been extensively validated. Fractional flow reserve (FFR) and the index of microvascular resistance (IMR) are well-supported by evidence for pressure wire-based assessment (17–19). However, recent advancements in functional assessment have introduced promising alternatives that eliminate the need for pressure wires. Kwon et al. were the first to validate and correlate the Murray law-based quantitative flow ratio (μFR) using a single angiographic view to calculate the epicardial flow ratio without requiring pressure wires or pharmacologically induced hyperemia (20). Similarly, non-hyperemic angiography-derived AMR has demonstrated prognostic performance comparable to invasive microvascular resistance, with high diagnostic accuracy when exceeding 250 mmHg*s/m, emerging as a practical computational tool for CMD evaluation and CFVR assessment (21–23).

Periprocedural myocardial injury (pMI) is a frequent occurrence in the context of coronary artery disease, particularly following PCI, with reported incidence rates ranging widely from 5% to 30%, depending on the definition used and the patient population studied (1). This significant variability in incidence stems from several factors, including differences in biomarker thresholds for diagnosis, the use of varying troponin assays with distinct sensitivities, and inconsistencies in the criteria applied to differentiate myocardial injury from infarction (24,25). Additionally, procedural variables such as lesion complexity, patient comorbidities, and operator technique can influence the likelihood of pMI (26).

This study highlights the strong link between CMD and pMI, as patients with pMI showed significant post-PCI impairments in AMR and CFVR. The significant post PCI increase in AMR observed in the pMI group (p <0.01) aligns with prior research indicating that elevated microvascular resistance after PCI correlates with myocardial injury. In the absence of side branch occlusions or flow-limiting dissections at the stent site, it is plausible that structural and functional microcirculatory impairment— potentially due to embolization of plaque debris and thrombotic aggregates—contributes to myocardial injury. The post PCI decrease in CFVR **(Table 3)** was also a marker of pMI, despite all patients having TIMI III flow based on visual estimation.

The mechanisms underlying pMI are multifactorial, involving both microvascular and procedural elements. Our study confirms that increased microvascular resistance following PCI correlates with higher rates of myocardial injury, consistent with prior research (27,28). A smaller study (n=50) by Mangiacapra et al. (29) previously demonstrated that a post-PCI wire-based index of microvascular resistance (IMR) >38 strongly predicts type 4a myocardial infarction. Several procedural factors have been implicated in the development of microvascular dysfunction and myocardial injury.

Distal embolization of plaque debris and thrombotic material is a well-recognized contributor to pMI (30). The significant increase in post PCI AMR observed in our cohort supports this concept, as microvascular obstruction due to embolized material likely leads to increased resistance and impaired perfusion. This is consistent with previous studies demonstrating that distal embolization results in an inflammatory cascade, further exacerbating microvascular resistance and limiting post-procedural coronary flow (31).

Mechanical factors such as balloon overexpansion and prolonged vessel manipulation have also been implicated in microvascular dysfunction (32). Excessive mechanical stress may lead to microvascular spasm, endothelial injury, and increased microvascular permeability, all of which contribute to increased resistance and impaired myocardial perfusion. This is rational, but we could not find any difference in pre or post dilatation balloon diameter comparing no pMI vs pMI group **(Table 3).**

The interplay between pre procedure microvascular dysfunction and procedural stress remains an area of active investigation. Some studies suggest that microvascular dysfunction predisposes patients to pMI by limiting the adaptive capacity of the microcirculation to accommodate increased blood flow following PCI (33,34). In contrast, as shown in **Table 3**, our findings indicate that patients who developed pMI had less severe microvascular dysfunction at baseline compared to those without pMI, suggesting that procedural factors—rather than pre-existing dysfunction—may be the primary drivers of microvascular impairment leading to myocardial injury. This finding challenges the traditional paradigm and suggests that microvascular damage induced by PCI itself may be the dominant mechanism.

Pharmacological interventions aimed at enhancing microvascular function post-angioplasty are under active investigation. Intensified dual antiplatelet therapy (DAPT), incorporating potent P2Y₁ ₂ inhibitors, has demonstrated efficacy in reducing platelet aggregation and subsequent microvascular obstruction PCI (35). Fanj J. et al. demonstrated that DAPT with Ticagrelor after successful PCI of a myocardial infarction improved CMD compared to Clopidogrel (36). Additionally, the intra-coronary administration of vasodilators such as papaverine has been explored to alleviate microvascular spasm and improve coronary blood flow, thereby enhancing microvascular perfusion (37). Glycoprotein IIb/IIIa inhibitors, including abciximab, eptifibatide, and tirofiban, have been employed during PCI to prevent platelet aggregation by blocking the final common pathway of platelet activation (38). These agents have shown potential in reducing thrombotic complications and improving microvascular outcomes, as indicated by The REVERSE-FLOW trial that tested bailout use of glycoprotein IIb/IIIa inhibitors in acute myocardial infarction patients with angiographic microvascular obstruction did not significantly reduce infarct size but decreased the extent of microvascular obstruction (39). These findings underscore the need for a balanced approach when considering pharmacological strategies to enhance microvascular function post-angioplasty, weighing the benefits of improved perfusion against the potential risks of adverse effects.

In summary, this study reinforces the critical role of the coronary microcirculation after successful PCI and underscores the importance of integrating microvascular assessment into routine clinical practice. Current data suggest that strategies to mitigate microvascular dysfunction, such as pharmacological approaches targeting endothelial health, could improve patient outcomes and reduce the burden of pMI (40, 41).

## 5 Limitations

The present study presents several limitations that must be acknowledged when interpreting the findings. First, the retrospective design inherently limits the ability to establish causality, and the reliance on electronic medical records may have introduced selection bias, as only patients with complete data were included. Additionally, the exclusion of patients with poor image quality or complex lesions, such as bifurcations requiring two-stent techniques, may have restricted the generalizability of the results to more straightforward PCI cases. The angiographic-derived microcirculatory indices used in this study, such as μFR and AMR, though innovative, are relatively new and may be less well-validated compared to invasive methods like pressure-wire assessments. Variability in software algorithms for these indices could have also influenced the results. Lastly, while the study demonstrated significant associations between post-PCI microvascular dysfunction and pMI, the mechanisms driving these relationships, such as distal embolization or microvascular spasm, were not directly assessed, necessitating further exploration.

## 6 Conclusion

In conclusion, this study highlights the critical role of microvascular dysfunction in the occurrence of pMI following elective PCI. These findings suggest that despite successful epicardial revascularization, patients with reduction in coronary flow (RVR) and increased microvascular resistance after PCI (and not at baseline) are more prone to have pMI. The use of non-invasive angiographic indices, such as RVR and AMR, provides valuable insight into microcirculatory function and could serve as important adjuncts in guiding PCI and assessing procedural success. However, more research is needed to further validate these indices and to develop therapeutic strategies specifically targeting microvasculature to improve post-PCI outcomes.

## 7 Impact on daily practice - Perspectives

The impact of this study on daily practice highlights the importance of routinely assessing microvascular function during PCI, using indices like μFR and AMR. By incorporating these non-invasive tools, clinicians can better identify patients at higher risk for pMI and adjust post-PCI management accordingly. This could lead to more personalized care, including targeted therapies to improve microvascular health and reduce complications. Ultimately, integrating microvascular assessment into standard practice may enhance PCI outcomes and reduce the incidence of pMI, improving overall patient prognosis.

## Data Availability

The data that support the findings of this study are available from the corresponding author upon reasonable request.

## Central Illustration: microvascular outcomes after uncomplicated elective PCI

(A) Schematic summary of the overall physiological changes observed in the cohort (n = 330) after successful elective percutaneous coronary intervention (PCI). Improvements were observed in coronary flow velocity ratio (CFVR) and quantitative flow ratio (μFR), whereas angiographic-derived microcirculatory resistance (AMR) and the prevalence of coronary microvascular dysfunction (CMD) also increased. (B) Prevalence of CMD (defined by AMR >250 mmHg*s/m) before and after PCI, stratified by the occurrence of periprocedural myocardial injury (pMI). In both groups, CMD significantly increased after PCI, particularly in patients with pMI (from 10.8% to 56.5%, P < 0.01), compared to those without pMI (from 19.1% to 39.7%, P < 0.01). (C) Post-PCI reduced velocity ratio (RVR) and the combined presence of CMD and RVR were both significantly more prevalent in patients with pMI than those without (RVR: 71.1% vs. 60.9%, P = 0.04; CMD + RVR: 50% vs. 19.1%, P < 0.01), highlighting their potential role in pMI pathophysiology.

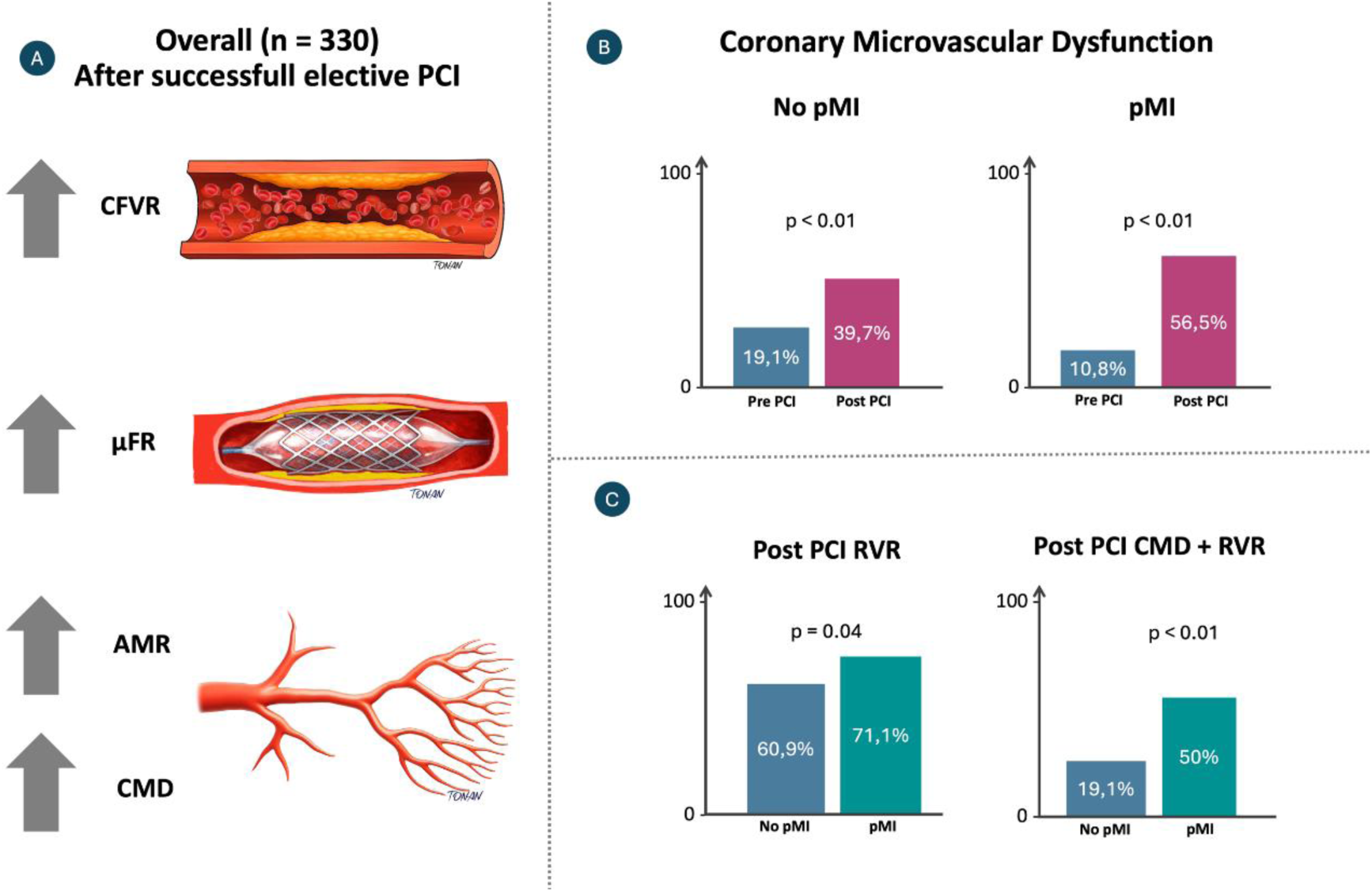

Caption: Coronary Artery Disease (CAD); Percutaneous Coronary Intervention (PCI); Periprocedural Myocardial Injury (pMI); Coronary Microvascular Dysfunction (CMD); Reduced Flow Velocity Ratio (RVR); Left Ventricular Ejection Fraction (LVEF); Creatinine Clearence (CrCl)

